# Cohort profile: The Westlake BioBank for Chinese (WBBC) pilot cohort: a prospective study for the late adolescence

**DOI:** 10.1101/2020.12.16.20248291

**Authors:** Xiao-Wei Zhu, Ke-Qi Liu, Ping-Yu Wang, Jun-Quan Liu, Jin-Yang Chen, Xue-Jin Xu, Jin-Jian Xu, Mo-Chang Qiu, Yi Sun, Chun Liu, Wei-Yang Bai, Pian-Pian Zhao, Jiangwei Xia, Si-Rui Gai, Peng-Lin Guan, Yu Qian, Pei-Kuan Cong, Shu-Yang Xie, Bin Wang, Hou-Feng Zheng

## Abstract

**Purpose:** The Westlake BioBank for Chinese (WBBC) pilot cohort is a population-based prospective study with its major purpose to better understand the effect of genetic and environmental factors on growth and development from adolescents to adults. The dataset comprises a wide range of demographics, anthropometric measures, physical activity, sleep quality, age at menarche and bone mineral density.

**Participants:** A total of 14,726 participants (4,751 males and 9975 females) aged 14 to 25 years were recruited and the baseline survey was carried out from 2017 to 2019. The pilot cohort contains rich range of information regarding of demographics, anthropometric measurements, blood pressure and heart rate, lifestyle and sleep patterns, biological, clinical and health outcomes.

**Findings to date:** There were most of the participants were Chinese Han ethnic (97.78%), and more than 60% of them were originally from rural areas. For anthropometry measurements, the mean height and weight were 172.92 cm and 65.81 kg for men, and 160.13 cm and 52.85 kg for women; the mean waist, hip and thigh circumference were 75.51cm, 90.75cm and 51.56 cm in men, and 71.67cm, 89.75cm and 51.68 among women. Results indicated that the prevalence of underweight in female was much higher than male, but the prevalence of overweight and obesity in female was lower than male. The mean of serum 25(OH)D level in the 14,726 young participants was 22.39 ng/ml, and male had higher level of serum 25(OH)D than female, overall, 33.47% of the participants had vitamin D deficiency and even more participants suffered from vitamin D insufficiency (58.17%). The proportion of deficiency in women was much higher than that in men (41.83 vs. 16.35%).

**Future plans:** WBBC is designed as a prospective cohort study and provides a unique and rich data set analyzing health trajectories of Chinese adolescents to young adults.

## Introduction

The initial goal of setting up this cohort is trying to bring up more attentions to the skeletal health in adolescence and adults, and thus to provide strategy in prevention of osteoporosis in the elderly. Osteoporosis, a disease of increased bone fragility, is a systemic osteopathy characterized by a decrease in bone density/quality and the destruction of bone microstructure caused by genetic and environmental factors^1^. Bone mineral density (BMD), the bone mineral content (BMC) in bone tissue, is recognized as the most important predictor of osteoporosis. Currently, approximately 200 million people worldwide suffer from osteoporosis, and 83.9 million of which are in China ^2^. As the trend of global aging is becoming more and more obvious, more attention should be paid to the bone health not only in the elderly, but also in adolescence and young adults. Population-based studies have shown that roughly half of the boys and one-third of the girls would undergo a fracture by age of 18 and 1/5 would have two or more fractures^3 4^. Epidemiologic studies have shown that a 10% increase in peak bone mass (PBM) at the population level reduces the risk of fracture later in life by 50%^5^. In fact, bone mass attained in early life was considered to be the most important modifiable determinant of lifelong skeletal health. A longitudinal data have shown that more than 94% of BMD was acquired at the age of sixteen^6^ and approximately 40% to 60% of adult bone mass was accrued during the adolescent years in both women and men^7^.

Any condition interfering with optimal peak bone mass accrual can, therefore, increase fracture risk later in life, adolescence is critical period for skeletal mineralization. The bone mass gain during adolescence is influenced by multiple factors, including genetic factors, ethnicity, and environmental factors, such as alcohol intake, cigarette smoking, physical activity, endocrine status (e.g. vitamin D), diet (calcium and protein intake) and other factors^5 8-13^. As environmental and behavioral factors account for 20% to 40% of adult peak bone mass^14 15^, the early identification of the factors associated with poor bone health and the provision of reliable counseling might help children and teenagers take action to maximize BMD before their PBM was completed.

Overweight and obesity are a major public health problem^16^. Elevated body mass index (BMI) in adolescence had been associated with several obesity-related morbidities in adult life, such as diabetes, metabolic syndromes and some types of cancer^17^. And obesity in adolescence conferred very high risks for obesity in adults^18^; 70% of overweight adolescents had one or more concomitant conditions such as high blood pressures and fasting insulin, which were also risk factors for cardiovascular disease, and 23% of those accompanied with three or more concomitant conditions^19^. Following rapid economic development since the 1980s, China experiences a rapidly increasing of overweight and obesity among children and adolescents^20 21^. In 2019, a cross-sectional study^22^ found that the prevalence of overweight in college students (aged 18-26 years) was 8.0%, and the prevalence of obesity was 3.5%. A recent study from 12 provinces in China showed that the prevalence of overweight and obesity were 14.0% and 10.5% in boys, and 9.7% and 7.1% in girls, respectively^23^.

Vitamin D deficiency is becoming a public health problem in both developed and developing countries^24 25^. Besides its effect on musculoskeletal system, Vitamin D showed pleiotropic effect on human health, such as cardiovascular diseases^26^, common infectious diseases^27^ and autoimmune diseases^28^. Serum 25(OH)D is a good indicator of vitamin D storage and is an optimal method of assessing vitamin D levels^24^. According to the Endocrine Society clinical practice guidelines, vitamin D levels were defined as deficiency [25(OH)D<20 ng/mL], insufficiency [25(OH)D: 20-29 ng/mL)] and sufficiency [25(OH)D≥30 ng/mL] respectively^29^. Many people in central and western Europe had vitamin D concentration of 11–20 ng/mL in winter ^30^. Studies from other countries, including Canada^31^, Japan^32^, Australia^33^ and Iran^34^, presented similar situations, with high prevalence of vitamin D insufficiency in different ethnicities. A study in north China found that more than 40% of adolescent girls had Vitamin D-deficiency in the winter^35^. Another study in Shanghai showed that more than one-third newborns had plasma 25(OH)D less than 20 ng/mL^36^. Even in Hong Kong (latitude 22° north), 72% of young adults were reported to have vitamin D deficiency^37^.

The overall goal of the Westlake Biobank for Chinese (WBBC) pilot cohort is to recruit individuals at their late adolescence/young adulthood. The biological samples such as whole blood, serum, urine and faeces were collected, genomic DNA were extracted and the DNA sequence information were acquired through sequencing technique. A long questionnaire with questions concerning the environmental factors such as nutrition, sleep quality, physical activity, medication etc was provided. These data will help us to understand the association between the genetics, environmental factors, microbiome and health statue of adolescence population. With a broad range of phenotype collection on many aspects of participants’ daily life, a wide range of scientific questions could be addressed. The main purpose of this particular paper is to profile the cohort, therefore, only limited findings were described, such as: (1) what are the prevalence of underweight, overweight, obesity and vitamin D deficiency in Chinese late adolescence? What is the reference value of serum vitamin D level in the young people? (2) what is the difference between male and female in term of height, weight, blood pressure, lifestyle and bone health in the young people.

## Cohort description

### Sampling Design

The Westlake Biobank for Chinese (WBBC) pilot study was collected in three main regions in China (Hanghzou city of Zhejiang province, Shangrao city of Jiangxi province and Yantai city of Shandong province), but the participants covered all around the country (Table 1 and Figure 1). The baseline survey was carried out from 2017 to 2019. The target population was young people aged 14–25 years who were college students and available for follow-up studies. In the first phase of baseline (WBBC pilot 1), the participants were recruited from two colleges at Zhejiang province and Jiangxi province in Southeast China from September 2017 to March 2018 (Figure 2), and 1,258 and 2,769 participants were from Zhejiang and Jiangxi provinces, respectively, and 1,263 participants were from other 25 provinces of China (Table 1). From September 2018 to December 2018, the second phase of WBBC pilot project was initiated (WBBC pilot 2), the participants were recruited at the same college in Jiangxi province and a college in Shandong province in Northeast China (Figure 2). There were 2,920 participants from Shandong province, 2,032 participants from Jiangxi province and 1,306 participants from other 28 provinces of China in WBBC pilot 2 (Table 1). From September 2019, the WBBC pilot project phase 3 (WBBC pilot 3) recruited participants from the same college in Jiangxi province (Figure 2), most of the participants (2,504) of WBBC pilot 3 were from Jiangxi province and 6,74 participants were from other 26 provinces of China (Table 1). All participants provided their Chinese unique national identity (ID) number for unique reference at the health examination center in the campus. The inclusive criteria were: a). All study participants signed the informed consent form before taking part in the survey. b). Participants should complete the physical examination, bone mineral density scan, blood test and questionnaires. And the exclusion criteria were: a). No informed consent. b). Participants missed most of the items in data collection. In WBBC pilot 3, the urine and feces of the participants were collected, therefore, participants taking antibiotics should be excluded.

**Table 1.** The participants of the cohort at baseline.

| Year | Phase | Area | Total | Sex |  |
| --- | --- | --- | --- | --- | --- |
|  |  |  |  | Male | Female |
| 2017 | WBBC pilot 1 | Zhejiang province | 1,258 | 460 | 798 |
|  |  | Jiangxi province | 2,769 | 862 | 1,907 |
|  |  | Other 25 provinces | 1,263 | 336 | 927 |
| 2018 | WBBC pilot 2 | Shandong province | 2,920 | 1,115 | 1,805 |
|  |  | Jiangxi province | 2,032 | 578 | 1,454 |
|  |  | Other 28 provinces | 1,306 | 438 | 868 |
| 2019 | WBBC pilot 3 | Jiangxi province | 2,504 | 761 | 1,743 |
|  |  | Other 26 provinces | 674 | 201 | 473 |
| Total |  |  | 14,726 | 4,751 | 9,975 |

**Figure 1.**
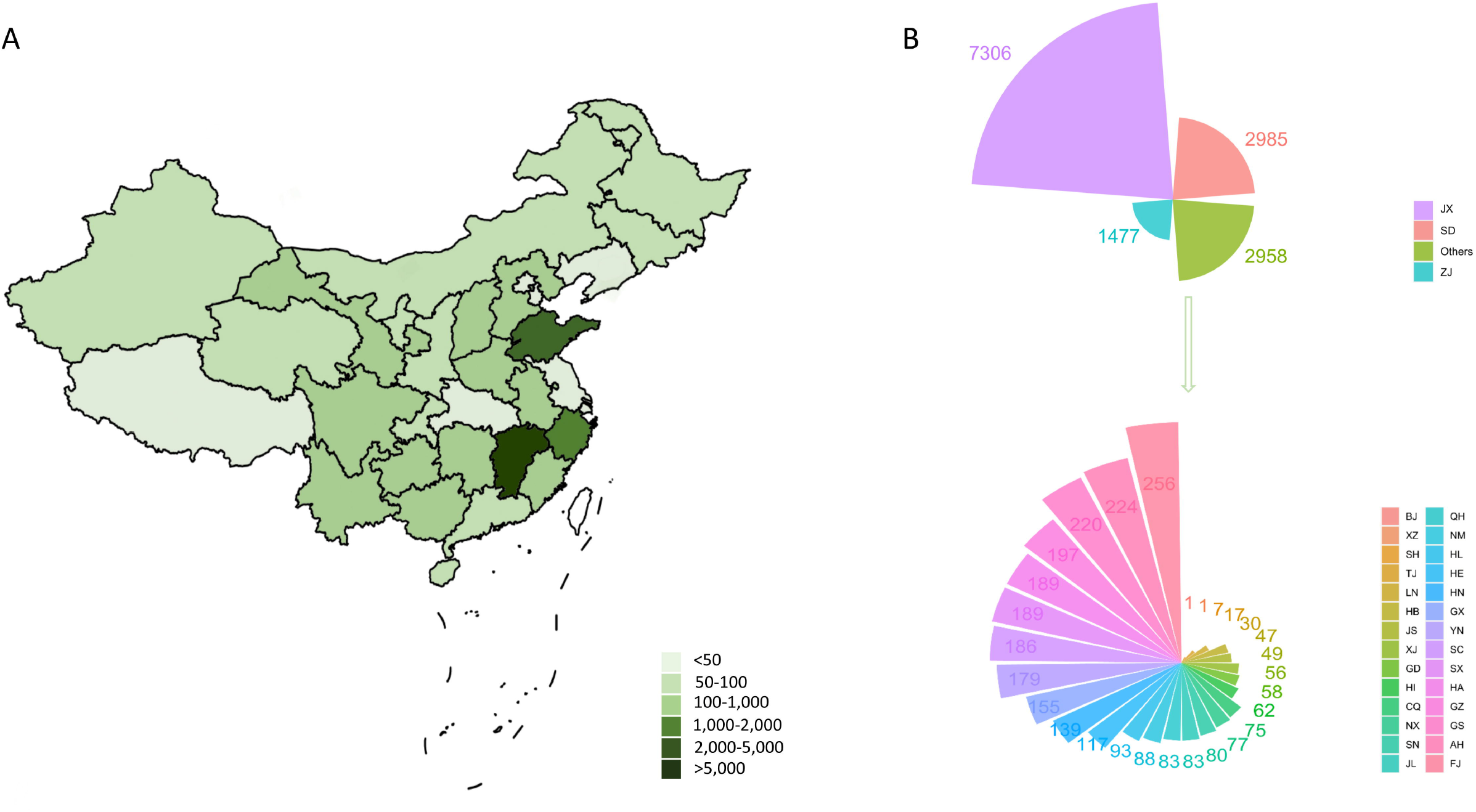
Maps showing the sources of the samples in the Westlake BioBank for Chinese (WBBC) pilot cohort. AH: Anhui province; BJ: Beijing; CQ: Chongqing; FJ: Fujian province; GD: Guangdong province; GS: Gansu province; GX: Guangxi Autonomous Region; GZ: Guizhou province; HA: Henan province; HB: Hubei province; HE: Hebei province; HI: Hainan province; HL: Heilongjiang province; HN: Hunan province; JL: Jilin province; JS: Jiangsu province; JX: Jiangxi province; LN: Liaoning province; NM: Neimeng Autonomous Region; NX: Ningxia Autonomous Region; QH: Qinghai province; SC: Sichuang province; SD: Shandong province; SH: Shanghai; SN: Shanxi province; SX: Shanxi province; TJ: Tianjin; XJ: Xinjiang Autonomous Region; XZ: Xizang Autonomous Region; YN: Yunnan province; ZJ: Zhejiang province.

**Figure 2.**
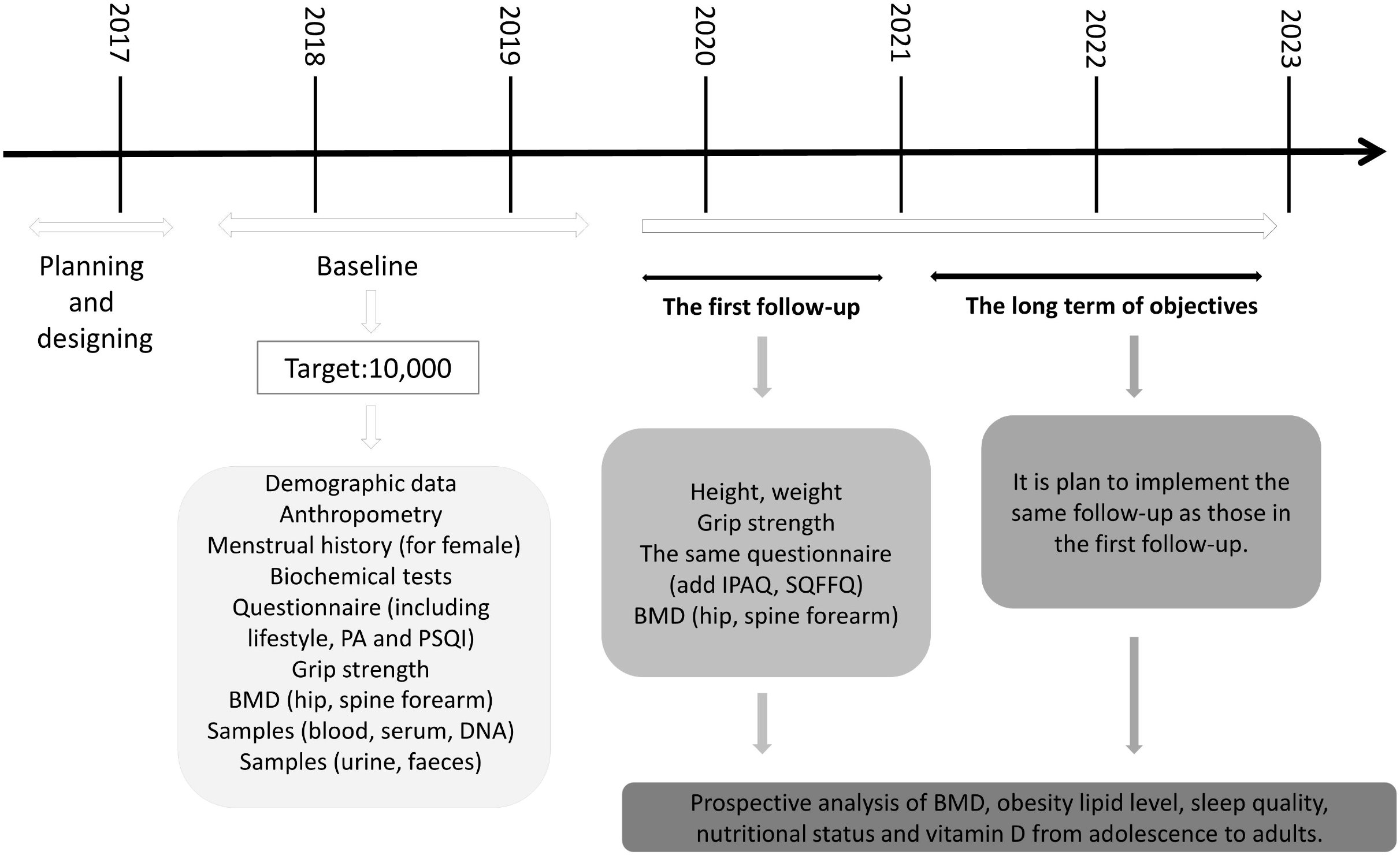
Data collection timeline. IPAQ: International Physical Activity Questionnaire; PA: Physical Activity; PSQI: Pittsburgh sleep quality index; SQFFQ: Semi-Quantitative Food Frequency Questionnaire.

### Data Collection Procedures

The WBBC pilot study is a multidisciplinary study and contains rich range of information regarding of demographics, anthropometric measurements, blood pressure and heart rate, lifestyle and sleep patterns, biological, clinical and health outcomes. Genomic data are available for 7,033 participants. Data and samples were collected via examinations, questionnaire and venipuncture (Table 2).

**Table 2.**
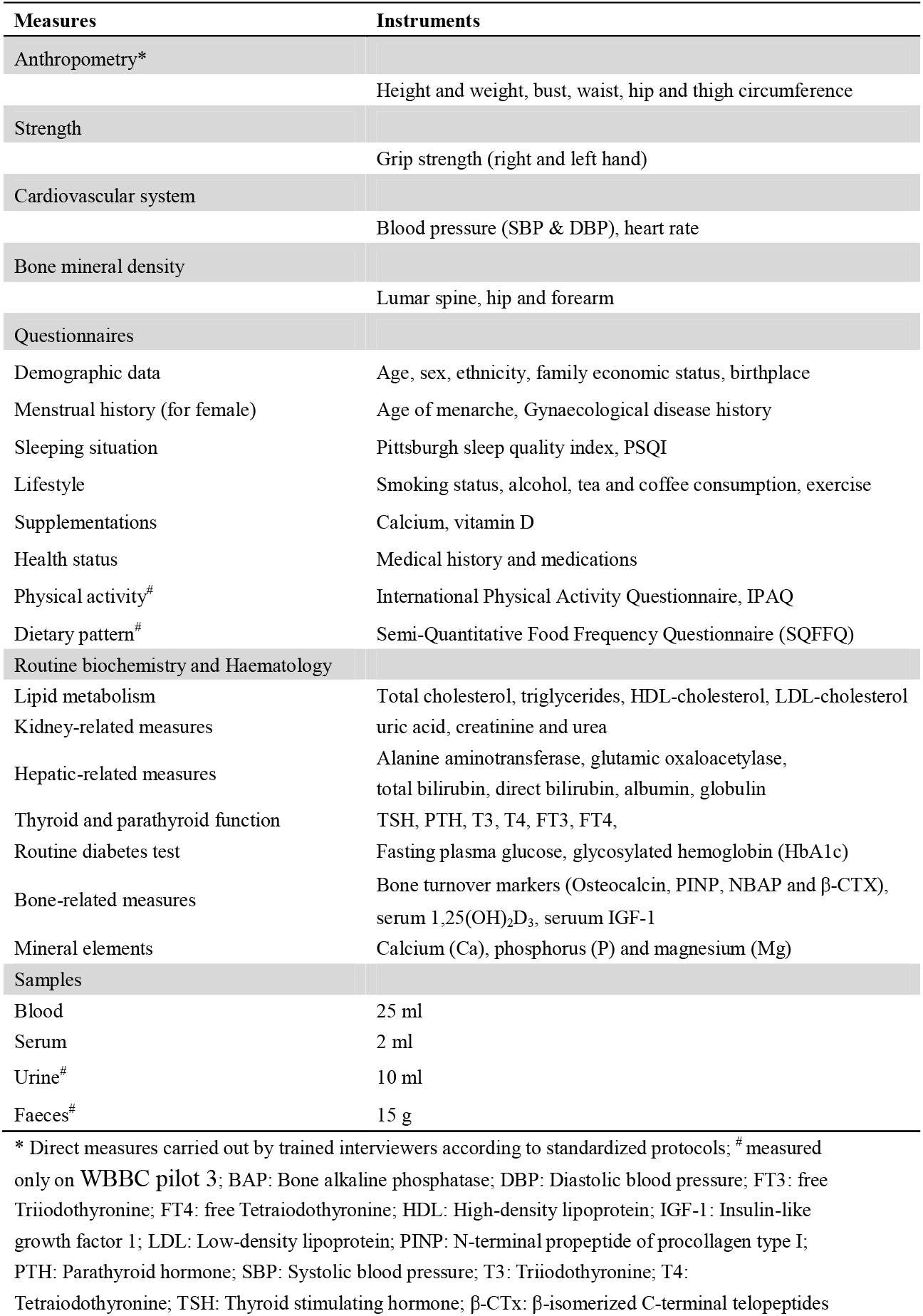
Summary of data collected in the Cohort.

| Measures | Instruments |
| --- | --- |
| Anthropometry* | Height and weight, bust, waist, hip and thigh circumference |
| Strength | Grip strength (right and left hand) |
| Cardiovascular system | Blood pressure (SBP & DBP), heart rate |
| Bone mineral density | Lumbar spine, hip and forearm |
| Questionnaires |  |
| Demographic data | Age, sex, ethnicity, family economic status, birthplace |
| Menstrual history (for female) | Age of menarche, Gynaecological disease history |
| Sleeping situation | Pittsburgh sleep quality index, PSQI |
| Lifestyle | Smoking status, alcohol, tea and coffee consumption, exercise |
| Supplementations | Calcium, vitamin D |
| Health status | Medical history and medications |
| Physical activity<sup>#</sup> | International Physical Activity Questionnaire, IPAQ |
| Dietary pattern<sup>#</sup> | Semi-Quantitative Food Frequency Questionnaire (SQFFQ) |
| Routine biochemistry and Haematology |  |
| Lipid metabolism | Total cholesterol, triglycerides, HDL-cholesterol, LDL-cholesterol |
| Kidney-related measures | uric acid, creatinine and urea |
| Hepatic-related measures | Alanine aminotransferase, glutamic oxaloacetylase, total bilirubin, direct bilirubin, albumin, globulin |
| Thyroid and parathyroid function | TSH, PTH, T3, T4, FT3, FT4, |
| Routine diabetes test | Fasting plasma glucose, glycosylated hemoglobin (HbA1c) |
| Bone-related measures | Bone turnover markers (Osteocalcin, PINP, NBAP and β-CTX), serum 1,25(OH)<sub>2</sub>D<sub>3</sub>, serum IGF-1 |
| Mineral elements | Calcium (Ca), phosphorus (P) and magnesium (Mg) |
| Samples |  |
| Blood | 25 ml |
| Serum | 2 ml |
| Urine<sup>#</sup> | 10 ml |
| Faeces<sup>#</sup> | 15 g |

\* Direct measures carried out by trained interviewers according to standardized protocols; <sup>#</sup> measured only on WBBC pilot 3; BAP: Bone alkaline phosphatase; DBP: Diastolic blood pressure; FT3: free Triiodothyronine; FT4: free Tetraiodothyronine; HDL: High-density lipoprotein; IGF-1: Insulin-like growth factor 1; LDL: Low-density lipoprotein; PINP: N-terminal propeptide of procollagen type I; PTH: Parathyroid hormone; SBP: Systolic blood pressure; T3: Triiodothyronine; T4: Tetraiodothyronine; TSH: Thyroid stimulating hormone; $\beta$ -CTX: $\beta$ -isomerized C-terminal telopeptides

#### Measurements of anthropometric parameters

Anthropometric data included height, body weight, bust, waist, hip and thigh circumference, resting blood pressure, heart rate and hand grip. Height was measured to the nearest 0.1 cm with participants’ light-weight clothes and shoes off; weight was measured to the nearest 0.01 kg with the weight scale (Ultrasonic surveying instrument, Beryl BYH01, China) calibrated daily before each series of measurements. Bust, waist, hip and thigh circumference were measured to the nearest 0.5 cm by using a measuring tape with the subject standing comfortably. Resting blood pressure and heart rate were measured on the left arm supported at heart-level sitting position using electronic sphygmomanometers (Yuwell YE660A, China). To ensure accurate data, the participants were asked to have rested for at least 5 minutes and have no excessive physical activity, tea or alcohol intake or smoking for at least one night. Using a handgrip dynamometer (CAMRY EH101, China), grip strength with both hands were tested for most of the participants in WBBC pilot 2 and WBBC pilot 3. In order to get more accurate results, the participants should make sure the arm that’s being tested was at a 90 degree angle at the elbow^38^ until the test was finished. Details of the methods and instruments used for measurements of anthropometric parameters were provided in Table 3.

**Table 3.** List of anthropometric collected and platforms for biochemical tests at baseline in the cohort.

| **Variables** | **Analysis method/platform used** |
| --- | --- |
| Height and weight | Ultrasonic surveying instrument, Beryl BYH01, China |
| Waist, hip and thigh circumference | Manufactured instrument (tape) |
| Resting blood pressure | Electronic sphygmomanometer, Yuwell YE660A, China |
| Heart rate | Electronic sphygmomanometer, Yuwell YE660A, China |
| Grip strength | Digital hand dynamometer, CAMRY EH101, China |
| Bone mineral density (BMD) | DXA, Discovery QDR 4500, Hologic Inc., Waltham, MA, USA |
| Blood routine (5 items) | SYSMEX 2100, Japan |
| Serum IGF-1 | CLIA, DPC immulite 2000, Siemens, Germany |
| Serum calcium, phosphorus | ARCHITECT C16000, Abbott, USA |
| Fasting blood glucose | Cobas c501, Roche, Switzerland |
| Bone turnover markers | ECLIA, Cobas e602, Roche, Switzerland |
| Serum 1,25(OH)<sub>2</sub>D<sub>3</sub> | LC-MS/MS, AB Sciex API 4000™, USA |
| Thyroid and parathyroid function | Chemical luminescence, ARCHITECT System i2000, USA |
| Lipid metabolism | ARCHITECT C16000, Abbott, USA |
| Kidney-related measures | ARCHITECT C16000, Abbott, USA |
| Hepatic-related measures | ARCHITECT C16000, Abbott, USA |

CLIA: chemiluminescent immunoassay; ECLIA: electrochemoluminescence immunoassay; LC-MS/MS: Liquid chromatography-tandem mass spectrometry.

#### Biochemistry assessment

Participants came to the examination center in each college in the morning with at least 8 h of overnight fasting, about 25 ml of venous blood samples were collected for routine blood measurements, biochemical indexes, DNA extraction and so on. Venous blood samples were collected using ethylenediamine tetraacetic acid dipotassium (EDTAK2) anticoagulation tube (3×5.0 ml) and vacuum tube without anticoagulation (2×5.0 ml). Serum and plasma samples were separated from whole blood through centrifugation for 10 min at the relative centrifugal force 3000 g (Figure 3). Serum samples were forwarded to test biochemical indexes that included serum 25(OH)D level, serum calcium level, fasting blood glucose, kidney function test, hepatic function test, blood lipids [triglyceride, cholesterol, low-density lipoprotein (LDL), high-density lipoprotein (HDL)], triiodothyronine (T3), thyroid and parathyroid function and bone turnover markers (Table 2). Details of the platforms used for biochemical analysis are provided in Table 3. We also reserved serum samples (2×0.5ml) for each participant at - 80□for future use. Figure 3 displays the detail of the flow diagram of blood separation and detection of main blood biochemical indexes.

**Figure 3.**
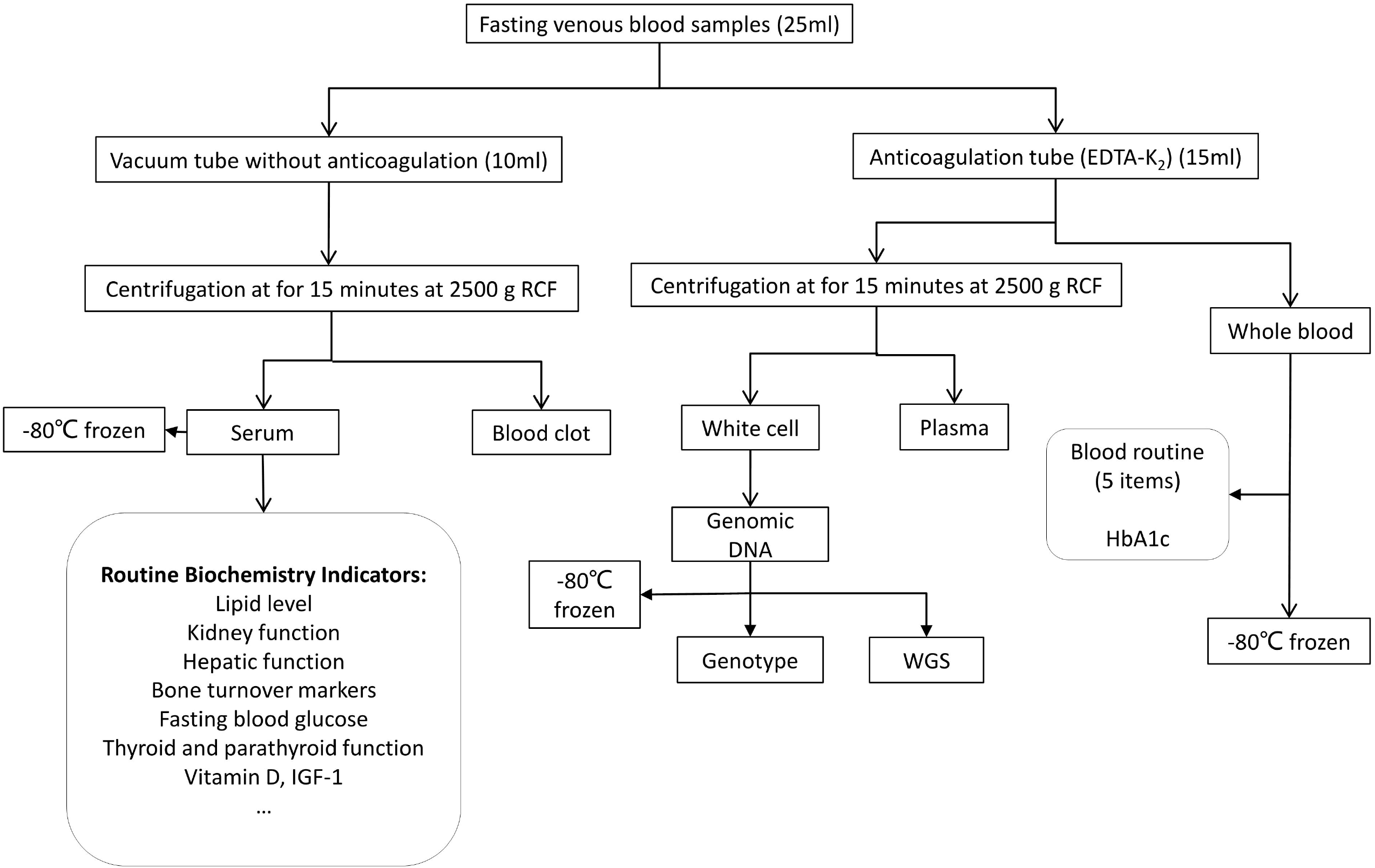
Flow diagram of main blood biochemical detection and blood conservation. RCF: relative centrifugal force; WGS: whole genome sequence.

#### Questionnaire-based assessments

Baseline data collection for participants included a self-completion questionnaire. In Table 2, a list of core questions within the aforementioned domains, was provided. The questionnaire included social and demographic measures data (e.g. age, sex, ethnicity, family economic status and born place), menstrual history (for female), lifestyle (e.g. physical activity, smoking status, alcohol, tea and coffee intake), additional supplement (e.g. calcium and vitamin D), health status and other information. Sleep duration and sleep quality were assessed by the Pittsburgh Sleep Quality Index (PSQI)^39^. This is composed of 19 questions which reflect seven major components, all seven components are then summed up to create a scale from 0–21 points.

#### Bone mineral density assessment

Bone mineral density (BMD) is expressed in terms of bone mass per cm^2^ (g/cm^2^) and were assessed by dual-energy X-ray absorptiometry^40^ (DXA, Discovery QDR 4500; Hologic Inc., Waltham, MA, USA). In WBBC pilot, the main sites for BMD evaluation were lumbar spine (L1–L4), femur (femoral neck and total hip) and distal third of the radius.

#### Whole Genome sequencing and genotyping

In the process of collecting fasting venous blood, 5-ml whole blood was used for isolation of genomic DNA. These genomic DNA were used for whole genome sequencing and genotyping (Figure 3). Whole genome sequencing was completed by NovaSeq 6000 system (Illumina Co., Ltd), and for now, 1,192 participants have been sequenced at mean depth of 14x, with highest depth of 65x. A Chinese specific reference panel will be constructed for imputation for Chinese population. Whole genome genotyping was completed with Infinium Asian Screening Array (ASA) (Illumina Co., Ltd), and 5,841 participants have been genotyped in approximately 700,000 SNPs.

#### Follow-up and outcome measures

We are seeking funding to follow the cohort to examine development and growth of the participants, and to investigate the effect of environmental factors on later outcomes. An important area of future research will focus on the development of bone mineral density and body weight from late adolescence to adulthood. Figure 2 shows the overall study plan. Follow-up surveys will be conducted according to the design of the subsequent research projects, the participants will be invited for re-survey with repeat interviews, including the questionnaire, anthropometric measurements, grip strength and bone mineral density collection as those used in the baseline stage and the data of nutritional status by food frequency questionnaire.

We have started a pilot follow-up study for WBBC pilot 2 since December 2019, and 1,303 participants had completed all examinations (Figure 2). The collected information included height, weight, grip strength and the updated questionnaire. Besides, we retested bone mineral density at spine (L1-L4), hip and distal third of the radius.

### Statistical analysis

To test differences in means and proportions between male and female, we used T-test and Chi-square tests for continuous and categorical variables, respectively. All variables were presented by unadjusted proportions for categorical variables and unadjusted means with standard deviations (SD) for continuous variables. The variables demonstrating a p-value of less than 0.05 were considered statistically significant. All statistical analyses were analyzed using Stata 12.0 software.

## Findings to date

This pilot cohort of WBBC is a large longitudinal survey conducted among adolescents and young adults in China. We surveyed 14,726 young people aged 14–25 years who were college students and available for completing follow-up studies. The baseline survey was carried out from 2017 to 2019, including WBBC pilot 1 (5,290 participants), WBBC pilot 2 (6,258 participants) and WBBC pilot 3 (3,178 participants).

We have several ongoing projects under WBBC pilot study. One of the most significant ongoing projects is the study of Chinese population structure. In WBBC pilot study, the data from 1,192 samples with whole genome sequencing and 5,841 samples with whole genome genotyping were available, collaborating with another research group in China, we finally achieved approximate ∼10,000 Chinese samples with whole genome data, covering 30 provincial region of China. In addition, WBBC pilot study will provide a haplotype reference panel to improve the imputation accuracy of Chinse GWAS study, since our previous study^41^ demonstrated that the existing reference panels, such as the 1000 Genome Phase3 panel^42^ and the HRC (Haplotype Reference Consortium) panel^43^, were not best fit for imputation for Chinese population, especially for the rare variant imputation.

Given the extensive range of data collection in the WBBC study, it is not feasible to present all the results, only limited findings were described in the present study. In summary, a total of 17,407 college students were invited, of whom, 14,983 (86.07 %) responded. After removing participants with missing data and invalid data, the final study included an effective sample size of 14,726 (84.60%) adolescents and young adults (with age from 14 to 25 years, and mean age at 18.5 years). Table 4 provides an overview of socio-demographic, anthropometry, cardiovascular system, lifestyle, grip strength and BMD characteristics of the WBBC pilot participants at baseline. Briefly, within the 14,726 sample, there were more females than males (67.74 vs. 32.26%), with mean age of 18.48 years for women and 18.55 years for men, respectively. Most of the participants were Chinese Han ethnic (97.78), and more than 60% of them were originally from rural areas (60.24% of males and 69.62% of females). For anthropometry measurements, the mean height and weight were 172.92 cm and 65.81 kg for men, and 160.13 cm and 52.85 kg for women; the mean waist, hip and thigh circumference were 75.51cm, 90.75cm and 51.56 cm in men, and 71.67cm, 89.75cm and 51.68 among women. The mean systolic blood pressure (SBP), diastolic blood pressure (DBP) and heart rate (HR) in participants were 113 mmHg, 71 mmHg and 86 beats/minute, respectively. In the cohort, only 5.49% of the participants were current smokers and 38.82% of them were regular drinkers. Regarding the current smoking status, there was significant difference between men and women (16.29 vs. 1.64%, p < 0.001). As for alcohol consumption, the proportion of current drinker in boys and girls were 62.03% and 29.19%, respectively, which is much higher in males (p < 0.001). The mean sleeping time estimated in women were higher than men (8.34 vs. 7.97 hours, p < 0.001). As for grip strength, the data collection was started from WBBC pilot 2, the mean of grip strength in boys were much higher than girls (grip-left: 36.66 vs. 27.38kg and grip-right: 39.90 vs. 29.76 kg, both p < 0.001).

**Table 4.** Basic characteristics of participants in baseline of WBBC pilot.

| Variables(unit) |  | Total | Sex |  | *P value |
| --- | --- | --- | --- | --- | --- |
|  |  |  | Male | Female |  |
| Socio-demographic |  |  |  |  |  |
| Age (years) | N<sup>a</sup> | 14,726 | 4,751 | 9,975 | 0.001 |
|  | M (SD)<sup>b</sup> | 18.50 (1.25) | 18.55 (1.21) | 18.48 (1.26) |  |
| Gender | N (%)<sup>c</sup> | 14,726 (100) | 4,751 (32.26) | 9,975 (67.74) | <0.001 |
| Ethnicity |  |  |  |  | 0.305 |
| *Han* | N (%) | 11,136 (97.78) | 3,642 (97.98) | 7,494 (97.68) |  |
| *Others* | N (%) | 253 (2.22) | 75 (2.12) | 178 (2.32) |  |
| Hukou status |  |  |  |  | <0.001 |
| *Rural* | N (%) | 5,951 (66.59) | 1,739 (60.24) | 4,212 (69.62) |  |
| *Urban* | N (%) | 2,986(33.41) | 1,148 (39.76) | 1,838 (30.38) |  |
| Anthropometry |  |  |  |  |  |
| Height (cm) | N | 14,277 | 4,588 | 9,689 | <0.001 |
|  | M (SD) | 164.24 (8.49) | 172.92 (6.57) | 160.13 (5.76) |  |
| Weight (kg) | N | 14,279 | 4,587 | 9,692 | <0.001 |
|  | M (SD) | 57.01 (12.32) | 65.81 (13.60) | 52.85 (9.07) |  |
| Waist (cm) | N | 12,396 | 3,905 | 8,491 | <0.001 |
|  | M (SD) | 71.67 (9.84) | 75.51 (11.07) | 69.90 (8.66) |  |
| Hip (cm) | N | 12,388 | 3,902 | 8,486 | <0.001 |
|  | M (SD) | 89.75 (7.28) | 90.75 (8.28) | 89.29 (6.73) |  |
| Thigh (cm) | N | 12,351 | 3,880 | 8,471 | 0.102 |
|  | M (SD) | 51.68 (5.69) | 51.56 (6.50) | 51.74 (5.27) |  |
| Cardiovascular system |  |  |  |  |  |
| SBP (mmHg) | N | 14,277 | 4,595 | 9,682 | <0.001 |
|  | M (SD) | 113 (12) | 121 (12) | 110 (11) |  |
| DBP (mmHg) | N | 14,276 | 4,595 | 9,681 | <0.001 |
|  | M (SD) | 71 (9) | 73 (9) | 70 (8) |  |
| Heart rate (beats/minute) | N | 14,295 | 4,599 | 9,696 | <0.001 |
|  | M (SD) | 86 (13) | 83 (13) | 87 (13) |  |
| Lifestyle |  |  |  |  |  |
| Smoking | N (%) | 435 (5.94) | 350 (16.29) | 85 (1.64) | <0.001 |
| Alcohol consumption | N (%) | 2,844 (38.82) | 1,333 (62.03) | 1,511 (29.19) | <0.001 |
| Sleeping time (hours) | N | 7,247 | 2,115 | 5,132 | <0.001 |
|  | M (SD) | 8.23 (1.37) | 7.97 (1.26) | 8.34 (1.40) |  |
| Grip strength (kg) |  |  |  |  |  |
| Left hand | N | 8,932 | 2,958 | 5,974 | <0.001 |
|  | M (SD) | 27.38 (8.77) | 36.66 (7.53) | 22.79 (4.82) |  |
| Right hand | N | 8,941 | 2,967 | 5,974 | <0.001 |
|  | M (SD) | 29.76 (9.66) | 39.90 (8.32) | 24.72 (5.35) |  |
| BMD (g/cm<sup>2</sup>) |  |  |  |  |  |
| Lumar spine | N | 10,154 | 3,293 | 6,861 | <0.001 |
|  | M (SD) | 0.910 (0.105) | 0.926 (0.112) | 0.903(0.100) |  |
| Total hip | N | 10,160 | 3,296 | 6,864 | <0.001 |
|  | M (SD) | 0.868 (0.127) | 0.932 (0.139) | 0.837 (0.108) |  |
| Femoral neck | N | 10,160 | 3,296 | 6,864 | <0.001 |
|  | M (SD) | 0.778 (0.125) | 0.846 (0.138) | 0.746 (0.104) |  |
| Forearm | N | 9,917 | 3,238 | 6,679 | <0.001 |
|  | M (SD) | 0.657 (0.059) | 0.705 (0.057) | 0.634 (0.453) |  |

<sup>a</sup> N: sample size; <sup>b</sup>M (SD): mean (standard deviation); <sup>c</sup>N (%): sample size (percentage); \* T-test and Chi-square tests for continuous and categorical variables respectively to refer the significant differences between males and females; DBP: Diastolic blood pressure; SBP: Systolic blood pressure.

Height and weight were measured using the standardized procedures. Body mass index (BMI) was calculated based on the formula: weight in kilograms divided by height in meters-squared (kg/m^2^). According to the Working Group on Obesity in China (WGOC)^44^, participants were defined as underweight (<□18.5 kg/m^2^), normal weight (18.5-23.9 kg/m^2^), overweight (24-27.9 kg/m^2^) and obese (≥□28 kg/m^2^). Therefore, the WBBC pilot study provided an overall prevalence of underweight, overweight and obesity among young participants of 24.26%, 11.50% and 5.03%, respectively (Table 5). The prevalence of underweight in female was much higher than male (26.41% vs. 19.70%, p < 0.0001), but the prevalence of overweight in female was much lower than male (9.03% vs. 16.71%, p < 0.0001) (Table 5), similarly, the prevalence of obesity in female (3.21%) was lower than in male (8.88%) (p < 0.0001) (Table 5). Waist circumference (WC) is good indicator of abdominal visceral fat distribution and is a strong predictor of diabetes mellitus and cardiovascular disease ^45^. It is meaningful to investigate the WC along with BMI among adolescents and young people. In WBBC pilot study, central obesity was defined as WC≥85 cm for men and as WC≥80 cm for women based on the recommendations of the WGOC ^44^. In the cohort of 12,396 participants, the prevalence of central obesity was 14.62%, which was higher in male than in female (19.10% vs. 12.55%, p < 0.0001) (Table 5).

**Table 5.** Distribution of BMI or waist by sex in participants in WBBC pilot, 2017-2019. N=14,264.

| Variables | Total | Male | Female | #P |
| --- | --- | --- | --- | --- |
| Underweight, N (%) | 3,460 (24.26%) | 903 (19.70%) | 2,557 (26.41%) | <0.0001 |
| Normal, N (%) | 8,446 (59.21%) | 2,507 (54.70%) | 5,939 (61.35%) | <0.0001 |
| Overweight, N (%) | 1,640 (11.50%) | 766 (16.71%) | 874 (9.03%) | <0.0001 |
| Obesity, N (%) | 718 (5.03%) | 407 (8.88%) | 311 (3.21%) | <0.0001 |
| Central Obesity<sup>δ</sup>, N (%) | 1,812 (14.62%) | 746 (19.10%) | 1,066 (12.55%) | <0.0001 |

<sup>δ</sup> Total sample size for Central Obesity was 12,396. <sup>#</sup> Chi-square tests was calculated between male and female.

In WBBC pilot study, the prevalence of underweight were high in both male (19.70 %) and female (26.41%), though the prevalence of moderate and severe underweight decreased from 9.2% in 1975 to 8.4% in 2016 in girls and from 14.8% in 1975 to 12.4% in 2016 in boys in the world^46^. This phenomenon may due to the modern aesthetics of human stature that thinness is preferred, especially in China^47 48^. Recently, a study involving 2,023 young female participants (70.5% subjects aged 20-25 years) from eight Chinese universities^48^ showed that 30.55% of the participants were underweight, and 57.39% of them would like to be much thinner, which would lead to more underweight individuals. Therefore, future studies should not only pay attention to the problem of obesity/overweight, but also to the underweight issue in young people.

Using simple anthropometric indices of body composition, such as BMI and WC, has been considered as a practical and valuable approach to the assessment of obesity for a long time. Waist-to-hip ratios (WHR), waist-to-height ratios (WHeR), a body shape index (ABSI)^49^ and body roundness index (BRI)^50^ were also as parameters of body fat and visceral adipose tissue volume. In WBBC pilot cohort, we had collected several anthropometric measures including height, weight, bust, waist, hip and thigh circumference and these data could help us examine the usefulness of these anthropometric parameters and identify the optimal cut-off of the parameters to evaluate overweight and obesity among adolescence and young people in future study. It is noteworthy that vitamin D deficiency in females was significantly worse than in males. This may due to the modern aesthetics of Chinese culture that paler skin is preferred, especially in females. A questionnaire related to vitamin D and sun exposure was conducted at a university in Nanjing, China and found that 75.0 % of the students lacked sun exposure because they would like to avoid dark skin, and most of students (82.7 %) used sun protection, and sunscreen use were more popular in females^51^, but it was reported that using the amount of sun cream recommended by World Health Organization exponentially suppressed vitamin D synthesis in the skin^52^. In WBBC pilot study, the mean serum 25(OH)D level was 22.39 ± 5.33 ng/ml for all the participants (male: 25.15 ng/mL and female: 21.05 ng/mL, p < 0.0001) (Table 6). Overall, 33.47% of the participants had vitamin D deficiency and even more participants suffered from vitamin D insufficiency (58.17%) (Table 6). In addition, the proportion of women with sufficient vitamin D was much lower than that of men (3.69 vs. 17.91%, p < 0.0001), while the proportion of deficiency in women was much higher than that in men (41.83 vs. 16.35%, p < 0.0001) (Table 6). Most of the participants (86.87%) preferred to stay indoors in spare time, the females were less willing to do exercise than males (53.68% vs 70.59%) (Table 7), and 44.58% of females hardly had outdoor activities, only 5.89% of females often had outdoor activities every week (Table 7). These results jointly suggested that the females had not enough sun exposure. Although food sources of vitamin D were not commonly recognized, only 10–20% of vitamin D in human bodies was obtained through food sources^53^. In WBBC pilot study, there was only 2.24% of the participants used vitamin D supplements (3.12% in male and 1.87% in female, p = 0.00098) (Table 6) and this was consistent with Zhou et al^51^, which found that only 5.6 % of the students used vitamin D supplements in a university of Nanjing, China.

**Table 6.** Distribution of Vitamin D level by sex in subjects in WBBC pilot, 2017-2018. N=11,370.

| Variables | Total | Male | Female | P |
| --- | --- | --- | --- | --- |
| Serum 1,25(OH)D, mean (SD) | 22.39 (5.33) | 25.15 (5.43) | 21.05 (4.73) | <0.0001<sup>η</sup> |
| Vitamin D deficient, N (%) | 3,806 (33.47%) | 610 (16.35%) | 3,196 (41.83%) | <0.0001<sup>#</sup> |
| Vitamin D insufficient, N (%) | 6,614 (58.17%) | 2,452 (65.74%) | 4,162 (54.48%) | <0.0001<sup>#</sup> |
| Vitamin D sufficient, N (%) | 950 (8.36%) | 668 (17.91%) | 282 (3.69%) | <0.0001<sup>#</sup> |
| Vitamin D supplementation<sup>δ</sup>, N (%) | 164 (2.24%) | 67 (3.12%) | 97 (1.87%) | 0.00098<sup>#</sup> |

<sup>δ</sup> The sample size for Vitamin D supplementation was 7,326. <sup>#</sup> Chi-square tests was calculated between male and female. <sup>η</sup>T-test was calculated between male and female.

**Table 7.** Participants’ general react about activity status in WBBC pilot, 2017-2018. N=7,326.^a^

|  | **Total** | **Male** | **Female** |
| --- | --- | --- | --- |
| What would you like to do in spare time? |  |  |  |
| *Stay indoors* | 6,364 (86.87%) | 1,754 (81.62%) | 4,610 (89.05%) |
| *Take part in some activities* | 962 (13.13%) | 395 (18.38%) | 567 (10.95%) |
| Do you do exercise initiatively? |  |  |  |
| *Yes* | 4,296 (58.64%) | 1,517 (70.59%) | 2,779 (53.68%) |
| *No* | 3,030 (41.36%) | 632 (29.41%) | 2,398 (46.32%) |
| How often do you have outdoor activities every week? |  |  |  |
| *Hardly* | 2,785 (38.02%) | 477 (22.20%) | 2,308 (44.58%) |
| *Occasionally* | 3,865 (52.76%) | 1,301 (60.54%) | 2,564 (49.53%) |
| *Often* | 676 (9.23%) | 371 (17.26%) | 305 (5.89%) |

<sup>a</sup> Data are showed as n (%) of participants.

## Strengths and limitations

The WBBC pilot study was designed to develop a longitudinal database on health-related factors for young people. Firstly, this was a comprehensive cohort study to pay attention to the health of adolescence and young adults, including over ∼14,000 participants (Table 1). Secondly, the WBBC pilot cohort is rich in longitudinal phenotypic data and archived biospecimens, including whole blood and serum as well as DNA samples. These resources will facilitate to digitize the information of the genomic, proteomic and metabolomic and microbiome of the participants, and further investigate the association of the information with the development of adolescence and young adults. Thirdly, this cohort has now accrued sufficient events for reliable analyses, and the major findings on adiposity, BMD, sleep quality, nutrition and growth for adolescence are expected over the next few years.

There are several limitations to this cohort design. First, although the participants covered all around the country, most of them were mainly from 3 provinces (Jiangxi, Shandong and Zhejiang), and the selection of participants was not random, but in a specific population (college students). Second, the participants were collected in 3 phases at different time points, and the phenotypic information was updated in the later phases, for example, the items in the questionnaire of different phases were not always the same, therefore, the phenotypic data were not always uniform in 3 phases.

## Data Availability

The data is not freely available in the public domain, but specific proposals and ideas for future collaboration would be very welcome.

## Collaboration

Participants have agreed to provide their pseudonymized data being made available to other approved researchers. The WBBC pilot study welcomes and offers global collaboration. The data is not freely available in the public domain, but specific proposals and ideas for future collaboration would be very welcome. Applicants for collaboration and more information are encouraged to contact Dr. Hou-Feng Zheng (Email address), the person in charge of this project.

## Acknowledgements

We gratefully acknowledge all the participants of this project, and thank all the people who helped us in the sample recruitment, including volunteers, laboratory technicians, nurses and clerical workers. We also thank the Westlake University Supercomputer Center for the facility support and technical assistance. In addition, we would like to thank Shanghai AvanTech BioSciences Co., Ltd for their support and assistance in sample storage and management.

## Contributors

Hou-Feng Zheng gained funding and conceived of the study, and all authors were involved in the design and collection data of the study. Xiao-Wei Zhu and Hou-Feng Zheng analyzed the data and wrote the paper and all authors commented on previous versions of the manuscript. All authors read and approved the final manuscript.

## Patient and public involvement

No patients were involved in the described linkage between existing registries providing an anonymous data set.

## Ethics Approval and Consent to Participate

The study protocol and informed consent procedure were approved by the Ethics Committees at Westlake University. All study participants signed the informed consent form before taking part in the study.

